# Digital Behavioural Therapy for Insomnia and its Effects on Depression and Anxiety: An Individual Participant Data Meta-Analysis

**DOI:** 10.64898/2026.08.05.26359652

**Authors:** Linh Cao, Christopher Gordon, Jack Anderson, Nathaniel Marshall

## Abstract

Insomnia is a transdiagnostic risk factor for depression and anxiety and frequently co-occurs with both conditions. Sleep restriction therapy (SRT) is considered a key active component of cognitive behavioural therapy for insomnia (CBT-I), is now delivered without therapist involvement via digital platforms such as SleepFix. Existing meta-analytic evidence suggests that digital behavioural therapy for insomnia (dBT-I) may improve anxiety and depression, but participant-level evidence remains limited. This individual participant data meta-analysis pooled data from two Australian randomised controlled trials (dBT-I n=220; control n=270; 78.3% female; mean age 66.0 years) to examine whether dBT-I, with SRT as the central component and delivered through the SleepFix program, reduces depressive and anxiety symptoms in adults with insomnia disorder, who were not specifically selected for anxiety and depression. We measured anxiety using the Generalised Anxiety Disorder 7-item scale and depression using the Patient Health Questionnaire-9 or Geriatric Depression Scale-15, with depression scores standardised to a common scale assuming a shared standard deviation of 4. We fitted linear mixed-effects models with random intercepts for participants and trials at Weeks 8 and 16, including baseline GAD-7 (mean 6.1, SD 4.8) in the anxiety model. dBT-I significantly reduced anxiety at Week 8 (mean difference −0.94 GAD-7 points, 95% CI −1.80 to −0.09, p=.030) and Week 16 (−0.94 GAD-7 points, 95% CI −1.86 to −0.02, p=.044), and depression at Week 8 (−0.40 SDs, 95% CI −0.66 to −0.14, p=.003) and Week 16 (−0.44 SDs, 95% CI −0.72 to −0.17, p=.002), with no evidence effects diminished between timepoints. However, the reductions were less than the smallest detectable difference for these questionnaires (i.e., 1 point). These findings support dBT-I as a scalable intervention with modest mental health benefits extending beyond insomnia.

## Introduction

Insomnia disorder, characterised by persistent difficulty initiating sleep, maintaining sleep, or waking too early despite adequate opportunity for sleep (1), is a recognised transdiagnostic risk factor for depression and anxiety, and these conditions frequently co-occur (2, 3). Untreated insomnia increases the risk of developing depression and predicts poorer treatment outcomes for existing mood disorders, whereas treating insomnia directly may reduce depressive symptoms and lower the risk of depression onset (4, 5).

Cognitive behavioural therapy for insomnia (CBT-I) is the recommended first-line treatment across international guidelines, and standalone sleep restriction therapy (SRT) might be the most effective component to improve chronic insomnia (6-9). Digital delivery of CBT-I has emerged as a scalable, low-cost way to broaden access, with programs centred on SRT to reach individuals who cannot access in-person care (10, 11), and digital platforms such as Sleepio, SHUTi, and SleepFix deliver this core behavioural component without requiring therapist involvement (11, 12). This paper explores SleepFix exclusively, which delivers digital SRT through personalised bedtimes and wake times informed by repeated sleep measurements, restricting time in bed to improve sleep efficiency.

Existing meta-analytic evidence suggests that digital therapy produces small-to-moderate reductions in depressive and anxiety symptoms alongside improved sleep, but the magnitude of effects varies considerably across studies, likely reflecting differences in study populations, intervention designs and digital platforms (13). These analyses pool study-level summary statistics rather than individual participant data, which are prone to aggregation bias and limit the ability to account for participant-level variation or to identify who benefits most from treatment (13). Individual participant data (IPD) meta-analysis addresses these limitations by pooling raw data from multiple trials, thereby increasing power, enabling appropriate clustering modelling, and improving flexibility in examining treatment effects (14).

However, despite growing evidence that digital insomnia interventions improve mental health outcomes, to our knowledge, no previous IPD meta-analysis has evaluated anxiety and depression outcomes across trials of a common digital SRT intervention. To address this gap, we pooled IPD from two completed randomised controlled trials of the SleepFix, a fully automated dBT-I platform developed at the Woolcock Institute of Medical Research, to estimate the effects of dBT-I on depressive and anxiety symptoms at Weeks 8 and 16. We hypothesised that dBT-I would produce greater reductions than control by Week 8, and that these reductions would be maintained at Week 16.

## Methods

### Study design and data sources

We conducted a secondary IPD meta-analysis of two parallel-group randomised controlled trials of SleepFix, a fully automated digital program based on SRT. ASTEROID (ANZCTR ACTRN12620001111910) compared SleepFix with a waitlist control in adults with chronic insomnia and measured depression with the PHQ-9. EXCEED (ANZCTR ACTRN12618001668291) compared SleepFix with sleep hygiene education in adults with insomnia and measured depression with the GDS-15. Both trials were designed and powered to detect changes in insomnia severity. Depression or anxiety were collected as secondary outcomes, making both trials suitable for this research question, but neither was individually powered to detect effects on these endpoints. Harmonisation was justified by both trials evaluating the same fully automated SleepFix intervention, recruiting adults who met insomnia disorder criteria, applying identical follow-up schedules with pre-specified outcomes at Weeks 8 and 16, and collecting the GAD-7 at matching time points within a common protocol framework. Both trials were also conducted under the same principal investigator, with compatible eligibility criteria and active or control conditions.

### Participants

Participants were adults with insomnia disorder enrolled in either trial. Across both datasets, 490 participants were included (dBT-I n=220; control n=270). Participants were predominantly female (78.3%), with a mean age of 66.0 years (SD 6.9) and a mean Insomnia Severity Index score of 17.9 (SD 4.2) at baseline, indicating moderately severe insomnia. As shown in Table 1, participants presented with mild anxiety symptoms at baseline (GAD-7 mean 6.1, SD 4.8). Baseline depression data were available only for EXCEED participants (GDS-15 mean 5.9, SD 3.7), as ASTEROID did not collect baseline PHQ-9. The sample therefore represents an Australian older adult population with clinically significant insomnia and low-to-mild comorbid psychological symptoms at baseline, rather than a population recruited for clinical depression or anxiety. Ethics approval was granted to each trial separately, and this secondary analysis used fully de-identified data within the scope of existing approvals.

**Table 1.** Baseline participant characteristics.

| **Trial** | **N** | **Age, years mean (SD)** | **Female n (%)** | **ISI mean (SD)** | **GAD-7 mean (SD)** | **Depression, GDS-15 mean (SD)** |
| --- | --- | --- | --- | --- | --- | --- |
| **ASTEROID** | 321 | 67.0 (5.8) | 223 (81.7%) | 17.4 (4.3) | 5.5 (4.4) | Not collected |
| **EXCEED** | 169 | 64.2 (8.4) | 123 (72.8%) | 18.9 (3.8) | 7.1 (5.1) | 5.9 (3.7) |
| **Overall** | 490 | 66.0 (6.9) | 346 (78.3%) | 17.9 (4.2) | 6.1 (4.8) | 5.9 (3.7) |

### Outcomes

Anxiety was assessed with the Generalised Anxiety Disorder 7-item scale (GAD-7; range 0-21) in both trials. Depression was measured with the Patient Health Questionnaire-9 (PHQ-9; ASTEROID; range 0-27) and the Geriatric Depression Scale-15 (GDS-15; EXCEED; range 0-15). Because the instruments measuring depression use different scales, raw depression scores could not be pooled directly. We therefore centred each score on its trial mean and divided by an assumed common standard deviation of 4 to produce a standardised depression score in standard deviation units. This approach assumes that the PHQ-9 and GDS-15 capture a common underlying construct of depressive symptom severity. We adopted a fixed value of 4 as a transparent approximation rather than an estimated parameter, as it is well supported by observed standard deviations ranging from 3.75 to 4.06 across both trials and all time points, and is consistent with values reported in clinical insomnia samples with comorbid depressive symptoms (4, 15, 16). Notably, the two trials’ depression instruments (PHQ-9 and GDS-15), despite differing scales and ranges, independently yielded highly similar observed standard deviations, which further supports the assumption that they capture a common underlying construct on a comparable scale of variability.

### Statistical analysis

Analyses followed intention-to-treat principles. We fitted separate linear mixed-effects models for anxiety and depression. Each model included fixed effects for group (dBT-I versus control), time (Week 8 and Week 16), and their interaction with random intercepts for participant and for trial. We included baseline GAD-7 (mean 6.1, SD 4.8) as a covariate in the anxiety model, while excluding a baseline depression covariate because ASTEROID did not collect baseline PHQ-9. Because baseline depression data were unavailable for ASTEROID, the depression models estimate between-group differences at follow-up rather than change from baseline. We reported the estimated marginal mean contrasts (dBT-I minus control) at Weeks 8 and 16, with 95% confidence intervals. We performed all data processing and statistical analyses using R (Version 4.5.2; R Core team, 2025) using the lme4 (version 2.0.1), lmerTest (version 3.2.1), and emmeans (version 2.0.2) packages. Author J.A., an experienced R coder, oversaw the R code by author L.C., and author N.M. independently reproduced the statistical results in SAS to confirm accuracy.

## Results

### Anxiety

dBT-I reduced GAD-7 scores relative to control at Week 8 (mean difference −0.94 points, 95% CI −1.80 to −0.09, p=.030). The effect remained evident at Week 16, with no meaningful change from Week 8; the marginal difference at Week 16 was −0.94 points (95% CI −1.86 to −0.02, p=.044). The difference between weeks 8 and 16 was not statistically significant (p = .999). Estimates are shown in Table 2.

**Table 2.** Mixed-effects model estimates for anxiety (GAD-7)

| **Contrast** | **Estimate** | **SE** | **95% CI** | **t** | **p** |
| --- | --- | --- | --- | --- | --- |
| dBT-I vs control, Week 8 | −0.94 | 0.43 | −1.80, −0.09 | −2.18 | .030 |
| dBT-I vs control, Week 16 | −0.94 | 0.47 | −1.86, −0.02 | −2.02 | .044 |

### Depression

dBT-I produced greater reductions in standardised depression scores relative to control at Week 8 (−0.40 SD, 95% CI −0.66 to −0.14, p=.003). The marginal difference at Week 16 was −0.44 SD (95% CI −0.72 to −0.17, p=.002). The difference between weeks 8 and 16 was not statistically significant (p = .717). Estimates are shown in Table 3.

**Table 3.** Mixed-effects model estimates for depression (standardised)

| **Contrast** | **Estimate** | **SE** | **95% CI** | **t** | **p** |
| --- | --- | --- | --- | --- | --- |
| dBT-I vs control, Week 8 | −0.40 | 0.13 | −0.66, −0.14 | −2.99 | .003 |
| dBT-I vs control, Week 16 | −0.44 | 0.14 | −0.72, −0.17 | −3.19 | .002 |

## Discussion

This IPD meta-analysis of two trials of the SleepFix program shows that dBT-I produces statistically significant reductions in both anxiety and depression relative to control. Both effects were present at Week 8 and remained evident at Week 16.

However, the magnitude of effects warrants careful interpretation. The reductions were less than the smallest detectable difference for these questionnaires (i.e., 1 point). These findings align with the broader literature indicating that digital therapy for insomnia produces small but consistent improvements in mood and anxiety (13, 15). Although the individual-level effects were modest, we interpreted them in the context of a scalable intervention that can be delivered at low cost with minimal therapist input. The persistence of effects through Week 16 is encouraging. Neither trajectory declined beyond Week 8, consistent with evidence that the behavioural mechanisms targeted by sleep restriction—sleep consolidation, reduced arousal, and improved daytime functioning—are maintained once established (9). This is consistent with the proposed mechanism by which SRT improves mood: by consolidating sleep and reducing arousal, the intervention may address the physiological substrate maintaining both insomnia and emotional dysregulation (9).

By analysing participant-level data from both trials, this IPD approach accounted for within-participant correlations across time and points for between-trial differences via random intercepts (14), which likely improved the precision of the effect estimates reported above relative to what a standard aggregate-data meta-analysis would have yielded. Although only two SleepFix trials were available, modelling clustering at both participant and trial levels allowed us to detect statistically significant effects despite the modest sample size, a result that may not have been achievable using study-level summary statistics alone (14).

Several limitations apply. First, the PHQ-9 and GDS-15 differ in item content and were validated in different populations, so some degree of measurement incomparability may remain despite standardisation (14). Second, the absence of baseline PHQ-9 data precluded adjustment for baseline depressive symptoms and may have reduced the precision of the estimated treatment effects. Third, the absence of participant blinding, inherent to behavioural trials, means a range of biased effects cannot be ruled out. Fourth, anxiety and depression outcomes relied on self-report questionnaires rather than clinician-administered assessment, which may be subject to reporting bias. Fifth, the analysis includes only two trials, so generalisability to other programs or to populations with more severe comorbidity is uncertain. Sixth, attrition was substantial: missing data at Week 16 were 56% overall for anxiety (62% dBT-I, 52% control) and 62% for depression (66% dBT-I, 59% control). Attrition was higher in the dBT-I arm than in the control arm across both trials and both outcomes. The mixed-effects models partially address this limitation by including all available observations under a missing-at-random assumption, but if missingness was related to the unobserved outcome itself, the treatment effect estimates may be biased in favour of dBT-I. Finally, harmonisable data beyond Week 16 were unavailable, so longer-term durability remains to be established.

In summary, this IPD meta-analysis provides evidence from two trials of SleepFix that dBT-I with SRT as the central component, although very modestly, produces a sustained, statistically significant reduction in anxiety and depression in adults with insomnia. The findings support dBT-I as a scalable intervention with low marginal costs and benefits extending beyond sleep. Future work should examine participant-level moderators of response and the relationship between engagement and mental outcomes, with a longer follow-up period.

## Acknowledgements

The authors thank the participants and research teams of the ASTEROID and EXCEED trials. No funding was specifically allocated to this secondary analysis.

## Author Contributions

L.C.: conceptualisation, data analysis, writing (original draft). C.G.: supervision, data access, intervention design. J.A.: statistical oversight and code review. N.M.: supervision, methodology, independent analysis, writing (review and editing). All authors approved the final manuscript.

## Data Availability Statement

The data that support the findings of this study are de-identified participant data from completed randomised controlled trials held by the Woolcock Institute of Medical Research. Data may be made available upon reasonable request to the corresponding author, subject to the terms of the original ethics approvals and approval by the relevant Human Research Ethics Committee.

## Ethics Approval Statement

The ASTEROID and EXCEED trials each received Human Research Ethics Committee approval. This secondary analysis used fully de-identified data within the scope of those existing approvals.

**Figure 1.**
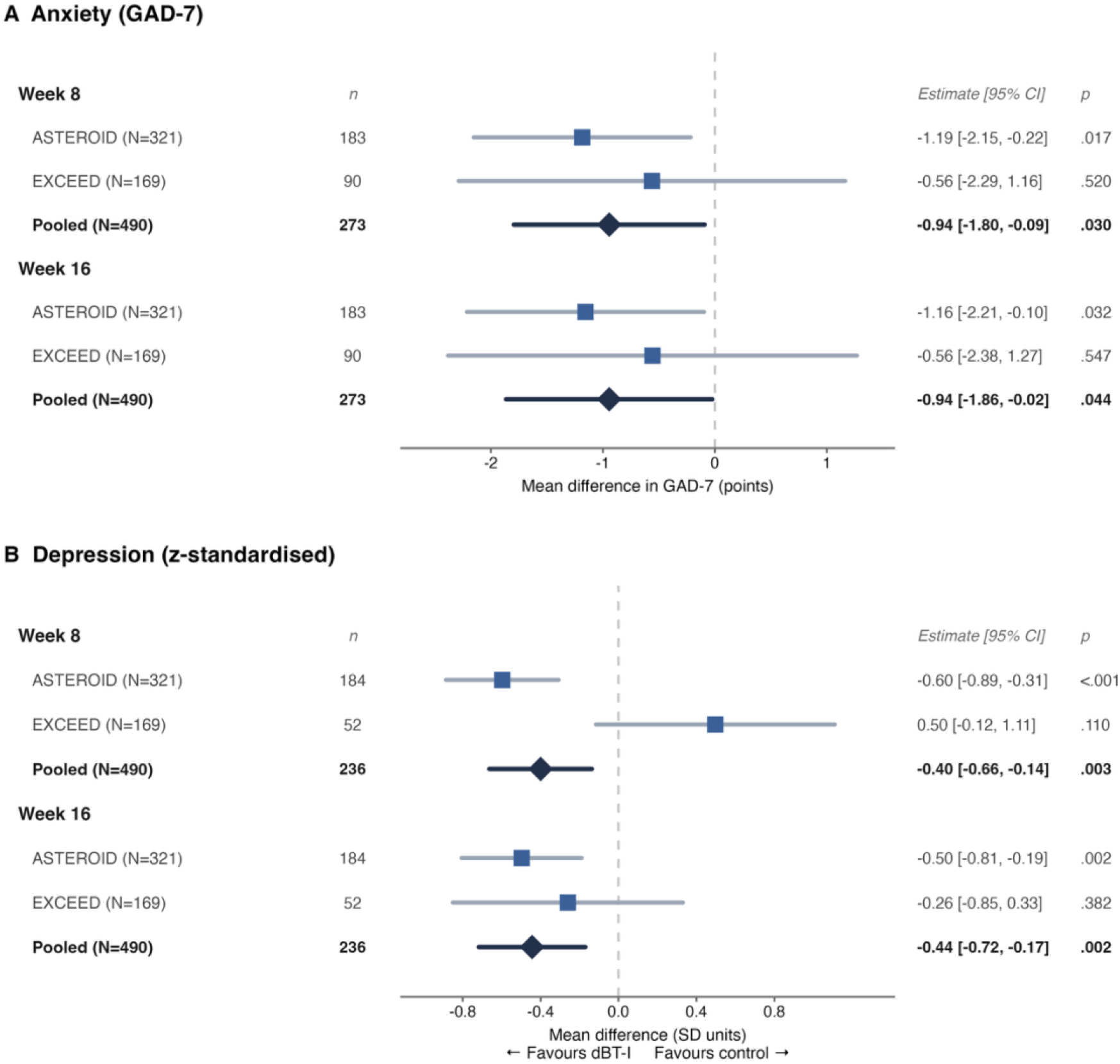
Estimated treatment effects (dBT-I vs control) on anxiety (A) and depression (B) at Weeks 8 and 16. Forest plots show study-specific and pooled estimates with 95% confidence intervals. Negative values favour dCBT-I. Abbreviations: dCBT-I, digital cognitive behavioural therapy for insomnia; GAD-7, Generalized Anxiety Disorder-7; PHQ-9, Patient Health Questionnaire-9.

